# The Effect of Integrative Reminiscence in Older Adults with Depression: A Systematic Review and Meta-Analysis

**DOI:** 10.1101/2025.04.07.25325360

**Authors:** Yuxuan Yang, Hongfei Li, Abdallah Abu Khait, Juliette Shellman

## Abstract

**Background:** Depressive symptoms are the most common comorbid psychiatric symptoms associated with older adults, given the physical, psychological, and emotional turmoil that most people experience. Integrative reminiscence, a type of intervention to combat these symptoms, emphasizes reflecting on one’s negative life events, resolving past conflicts, and reconciling the discrepancy between ideal and discovering personal meaning and worth in life. The review aimed to evaluate the efficacy of integrative reminiscence interventions in mitigating depressive symptoms among older adults.

**Methods:** A comprehensive literature search was conducted across four electronic databases: PubMed, CINAHL, PsycINFO, and Scopus. The search encompassed studies published in the English language. The risk of bias in randomized control and quasi-experimental studies was critically evaluated by the Cochrane ROB 2 tool, and the Cochrane ROBINS-I tool, respectively.

**Results:** A total of nine articles were included in the review. The meta-analysis showed a large and statistically significant effect of integrative reminiscence in reducing depression from post-baseline to baseline (Cohen’s *d* = 0.9608; *p* < .0001). The effects at 3-month follow-up or longer showed a similarly large effect size (Cohen’s *d* = 1.3159; *p* < 0.0001), indicating the effect of integrative reminiscence could be sustained over time for older adults with depressive symptoms.

**Conclusion:** This review demonstrates the significantly large effect of this intervention in alleviating depressive symptoms and provides robust empirical evidence for employing integrative reminiscence to reduce depressive symptoms in older adults.

## Introduction

According to the World Health Organization (2022), the number of older adults has rapidly increased in recent years due to improvements in health services. The proportion of those aged 60 and above will roughly increase from 12% to 22% in 2050. Globally, the adverse impact of aging on mental health is becoming more apparent as the incidence of depression among older adults increases exponentially. Depressive symptoms are among the most common comorbid psychiatric symptoms associated with older adults, given the physical, psychological, and emotional turmoil that most people experience (Baquero, 2015). Depressive symptoms in older adults can be characterized by distinct changes in mood, causing sadness or irritability (American Psychiatric Association, 2013). The level of depression among older adults can certainly progress from mild to severe levels (Petersen et al., 2017). Suicidal thoughts may eventually develop, which frequently come with various psycho-physiological changes like disturbances in appetite, sleep, and sexual urges.

Depression is often curable with early diagnosis and appropriate management. However, untreated minor and moderate depressive symptoms may progress into a major depressive disorder, leading to more suicide attempts and higher mortality rates (Nina, 2014; Oluboka et al., 2018; Petersen et al., 2017; Yang, 2017). Depressive symptoms are associated with a lower quality of life, greater disability in daily activities due to the increased progression of cognitive decline, a greater likelihood of nursing home placement, and an increased burden on caregivers (Nina, 2014; Youngsoon Yang, 2017). Older adults with depressive symptoms may experience the overgeneralized autobiographical memory phenomenon, which lies in the tendency of those older adults to reflect on negative memories more than positive ones (Hofer et al., 2017).

Attention has shifted toward non-pharmaceutical interventions, such as reminiscence therapy to mitigate depressive symptoms among older adults (Gonzalez et al., 2015; Meyer & O’Keefe, 2020). At all ages throughout the lifespan, most people can recall their past experiences and memories (Kovach, 1991). Older adults tend to retain vivid memories of the distant past, which they may be able to retrieve during the reminiscence process. Reminiscence is the natural process of recalling the past (Butler, 1963). Although it is a spontaneous process that happens to older adults, it can be used volitionally as a structured intervention to ameliorate mental disorders, particularly depression (Butler, 1963). A trained facilitator prepares meaningful topics that catalyze a planned reminiscence intervention focusing on older adults’ early childhood memories (Webster et al., 2010). The modality of a reminiscence intervention could be in either a group-based or individual-based format. Group reminiscence enables interaction among group members, while individual reminiscence serves as a platform that facilitates human-centered care (Van Bogaert et al., 2016).

Recent literature has acknowledged reminiscence’s positive impact and therapeutic merit as a non-pharmaceutical treatment for older adults (Aşiret & Kapucu, 2016; O’ Philbin et al., 2018). Many reviews have shown that reminiscence fulfills valuable functions for improving depressive symptoms (Huang et al., 2015; Irazoki et al., 2017; Woods et al., 2018). Reminiscence can help older adults increase self-acceptance and enable the resolution of past unresolved conflicts (Bohlmeijer et al., 2005; Huang et al., 2015; Shellman, 2016). This type of reminiscence is known as integrative reminiscence. Integrative reminiscence is more than the simple recollection of the past; it emphasizes reflecting on one’s negative life events, resolving past conflicts, and reconciling the discrepancy between ideal and discovering personal meaning and worth in life (Lieberman & Tobin, 1983; Watt & Cappeliez, 2000). Encountering negative life experiences can increase individuals’ susceptibility to depression. Interventions using integrative reminiscence focus on a constructive reevaluation of the meanings attributed to past self-defining events (Watt & Cappeliez, 2000). Through integrative reminiscence therapy, older adults can reintegrate negative life experiences, provide a sense of meaning in life, and achieve self-actualization, thereby improving depressive symptoms (Bohlmeijer et al., 2008).

Reminiscence was perceived as a unitary phenomenon until the development of a taxonomy. Watt and Wong (1991) introduced a taxonomy of reminiscence types: integrative, instrumental, transmissive, narrative, escapist, and obsessive reminiscence. Watt and Wong (1991) examined the relationship between various types of reminiscence and successful aging in older adults. Successful agers who exhibited better mental and physical health and adjustment showed significantly more integrative and instrumental reminiscence. The result invokes further exploration of the potential therapeutic utility of integrative reminiscence for depression.

However, a substantial number of clinical trials still employ non-specific reminiscence interventions, of which some approach reminiscence as a unitary phenomenon and ignore the differences within various reminiscence types (Yang et al., 2022). Consequently, a conspicuous lack of consistency is present in both the conceptualization and operation of reminiscence within the trials (Parker, 1995). The inconsistency in the findings of trials renders the therapeutic benefits of reminiscence inconclusive, which further diminishes the generalizability and replicability of these interventions (Watt & Cappeliez, 1995).

Given that reminiscence intervention is a general concept rather than a singular intervention, conducting a meta-analysis of the effectiveness of reminiscence interventions is inappropriate. The disparities between each type of reminiscence are remarkable, resulting in different types of reminiscence interventions that should be considered as different interventions. The combination of all reminiscence interventions, in turn, introduces statistical heterogeneity due to the variability in interventions and their ensuing clinical diversity (Haidich, 2014). Nevertheless, the existing meta-analysis conducted to investigate the efficacy of reminiscence on depression in older adults without considering the variety of reminiscence contributing to the efficacy of reminiscence remains inconclusive and less convincing (Liu et al., 2021; Tam et al., 2021). Therefore, the study aims to evaluate the efficacy of integrative reminiscence interventions in mitigating depressive symptoms among older adults. Moreover, the study aims to explore how the content of protocols influences the outcomes of integrative reminiscence intervention and provide the latest and valid evidence for developing a standardized intervention procedure.

## Methods

This systematic review and meta-analysis were conducted in accordance with the guidelines in the Preferred Reporting Items for Systematic Reviews and Meta-Analyses (PRISMA).

### Inclusion and Exclusion Criteria

The studies in this review were selected based on the following inclusion criteria: (a) studies involving older adults aged 60 and above; (b) experimental studies implementing integrative reminiscence as a primary intervention; (c) studies measuring depressive symptoms as an outcome; (d) studies published in peer-reviewed English-language journals up to August 2023. Exclusion criteria were: (a) not data-based studies (e.g., editorials, reviews, and commentary); (b) explicitly concentrating on subjects diagnosed with specific medical conditions (e.g., heart failure, cancer).

### Search Methods

A comprehensive literature search was conducted across four electronic databases: PubMed, CINAHL, PsycINFO, and Scopus. The search encompassed studies published in the English language up until August 2023. No other limitations were imposed on the publication year.

The keywords used in all the databases are “integrative reminiscence,” AND “depression,” AND “older adults,” and other related synonyms. The reference lists of the included studies were meticulously screened manually to identify any potentially eligible articles.

### Selection Process

After eliminating duplicate articles, the search results were extracted into an Excel spreadsheet. In the initial screening, the first and second authors independently screened the articles based on their titles and abstracts. Subsequently, the two authors reviewed the full text of the remaining literature and excluded studies that did not align with the inclusion and exclusion criteria.

### Data Extraction

The first and third authors performed data extraction using a self-developed, standardized data extraction form. The following information was extracted: author, year of publication, country, setting, study design, sample characteristics (e.g., sample size, mean age, proportion of women, and depression level), facilitators, details of the intervention (e.g., modality, length and number of sessions, memory triggers, and topics), details of the control, depression measurement instruments, and time points for outcome assessment. To calculate the effect measure related to depression outcomes, summary statistics (e.g., Mean, SD, F-statistics) were collected from the original articles upon data availability.

### Study Risk of Bias Assessments

Risk of bias assessments were conducted independently by the first and third authors. The risk of bias in randomized control and quasi-experimental studies was critically evaluated by the Cochrane ROB 2 tool (Higgins et al., 2019) and the Cochrane ROBINS-I tool (Sterne et al., 2016), respectively. Robvis created graphs to visually represent the results of the quality appraisal (McGuinness & Higgins, 2021).

### Synthesis Methods

In the meta-analysis, all data analysis and visualization were conducted using the R software (Lüdecke, 2018; R Core Team, 2023; Viechtbauer, 2010). The choice between fixed effect and random effect models is determined by the magnitude of heterogeneity observed across the selected studies, assessed by Cochran’s Q and I^2^. Specifically, when heterogeneity was low, we employed a fixed effect model, whereas in cases of high heterogeneity, a random effect model was used to account for potential variability.

Our primary outcome measure for meta-analysis was the effect size, with one effect size estimated per article included in our study. Since the selected articles were published in different years, the effect size calculations are not universal. Thus, it is unfair to perform the meta-analysis with different measures of effect size. In this meta-analysis, instead of extracting directly from the original article, the Standardized Mean Difference (SMD), namely Cohen’s d, is calculated for each article as the analysis effect size based on the following rule: (a) If both the mean and SD of pre-and post-test scores for both integrative reminiscence and control groups are available, the effect size is calculated through the standard mean difference in depression score reduction. (b) If the Mean of pre-and post-test scores is available, but the SD is not, it can be estimated by the square root of the mean square error (MSE) from ANOVA or ANCOVA. (c) If the Mean for the baseline and postbaseline scores is not available or if MSE is not available, the effect size is approximated by the F-statistics corresponding to the time-group interaction term from ANOVA or the F-statistics corresponding to the group term after adjusting for baseline depression score from ANCOVA. In this meta-analysis, the weight for aggregating the studies is decided by the sample size of each study instead of the inverse-variance approach, as a fact that the variance of the effect size could be relatively less robust since it depends on multiple causes like the SD of the test scores and the correlation of pre and post-tests, and the information is not always available.

In addition to the primary analysis, three subgroup analyses were conducted to explore potential differential effects: intervention group size, setting of the intervention, and type of control group. The group size was categorized as “small group form” for studies with four or fewer participants, including individual interventions, and “large group form” for those with more than four participants, with unreported sizes labeled as “Unknown.” Settings were classified as community or nursing homes, with one study in a veteran’s home included in the latter. Types of control groups were divided into active and waitlist controls. Additionally, we assessed the long-term effects of the intervention based on data availability.

To provide a comprehensive representation of the results, forest plots were generated to visualize the meta-analysis outcomes, facilitating the assessment of effect sizes and their confidence intervals across studies. Additionally, funnel plots were constructed to evaluate potential publication bias within the included studies, ensuring the robustness of our findings.

## Results

### Study Selection

The literature search and selection process are outlined in Figure 1. Initially, a search across four databases in July 2023 surfaced 163 articles, with one additional article sourced from manual reference list checks. Once 53 duplicates were removed, the remaining 110 articles were evaluated by their titles and abstracts. Subsequently, the remaining 55 full-text articles were reviewed based on exclusion criteria, of which 10 were retained for quality appraisal. One article was excluded at the synthesis stage due to insufficient data availability (only mean response is available without providing standard error) to calculate effect size (Sabir et al., 2016), leaving a total of nine articles for the meta-analysis (Bohlmeijer et al., 2009; Karimi et al., 2010; Meléndez Moral et al., 2015; Musavi et al., 2017; Pearson, 2006; Pinazo-Hernandis et al., 2022; Shellman et al., 2009; Watt & Cappeliez, 2000; Wu, 2011).

**Figure 1.**
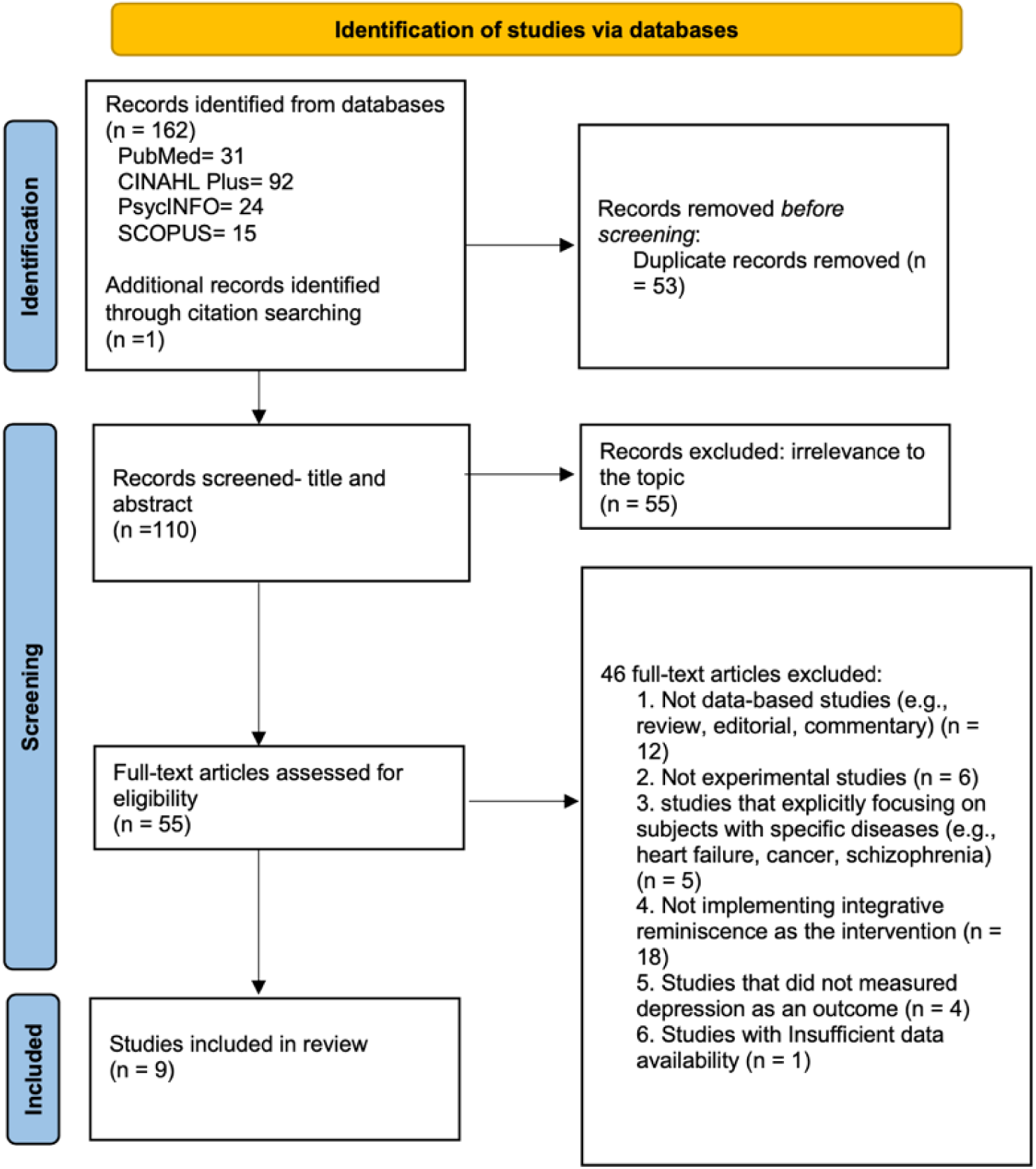
PRISMA 2020 Flow Diagram.

### Study Characteristics

Table 1 summarizes the characteristics of the included studies in each study. The nine studies included 396 older adults aged 60 and above. The sample sizes range from 26 to 108, and the mean age varies from 63.8 to 87.7, with an overall mean of 76.4. Female participants predominated in 66.7% of the reviewed studies, with two exclusively involving female participants and one recruiting only male participants. Geographically, the studies are globally distributed, with two each in the U.S. and Iran and the remaining disseminated across the Netherlands, Spain, Tunisia, Canada, and Taiwan. Five of the nine trials employed a randomized control trial (RCT) methodology, and the remaining 4 were quasi-experimental designs with control groups. The majority (n = 8) implemented the integrative reminiscence intervention in a group format, in contrast to the only study that deployed individual approaches. Variations were also observed in the frequencies and durations of the integrative reminiscence interventions, with sessions numbering between 6 and 20 and each spanning 45 to 120 minutes. All studies included in this review had established a treatment protocol that introduced the content and the discussion topics in the integrative reminiscence interventions.

**Table 1.**
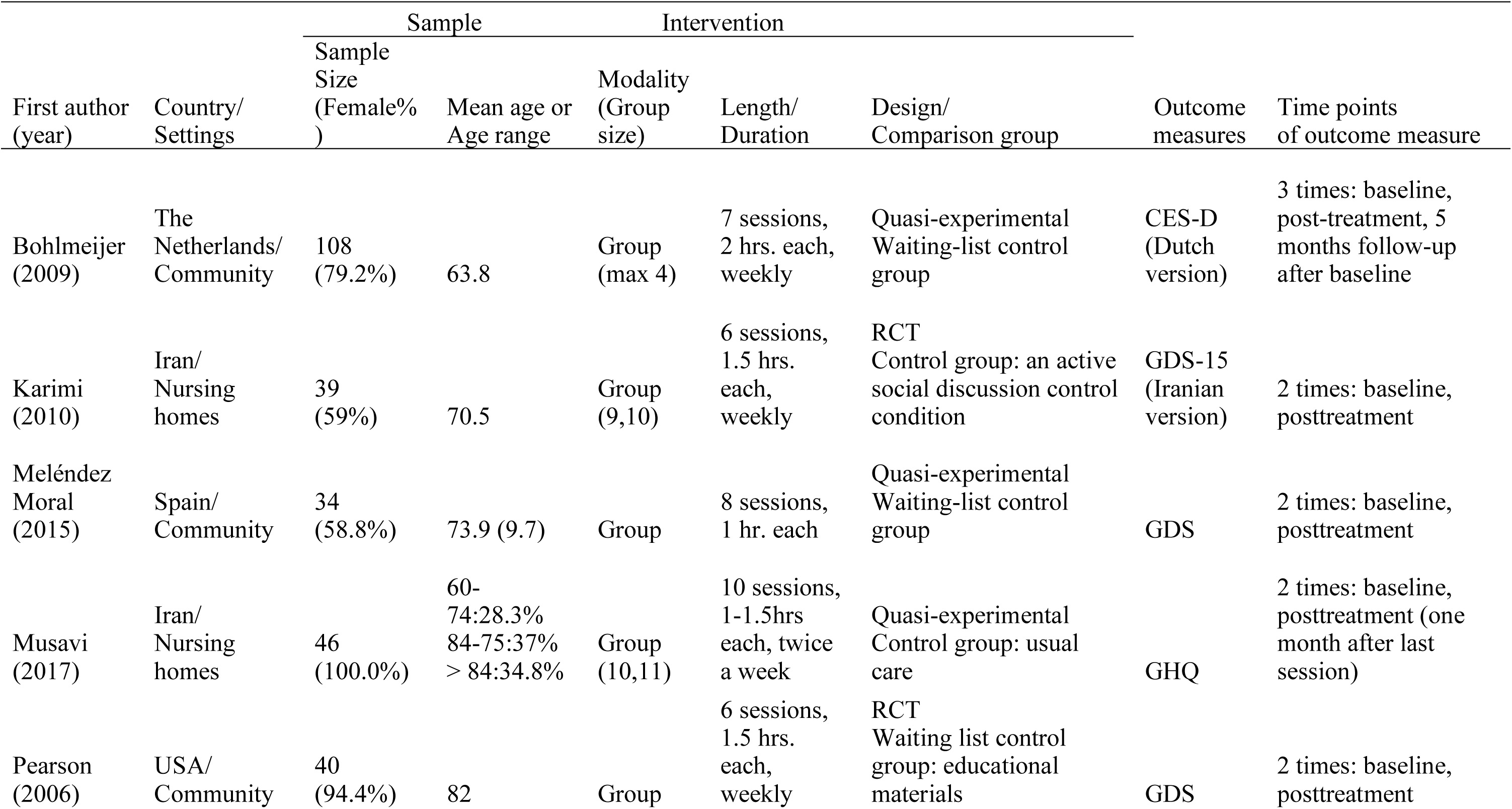
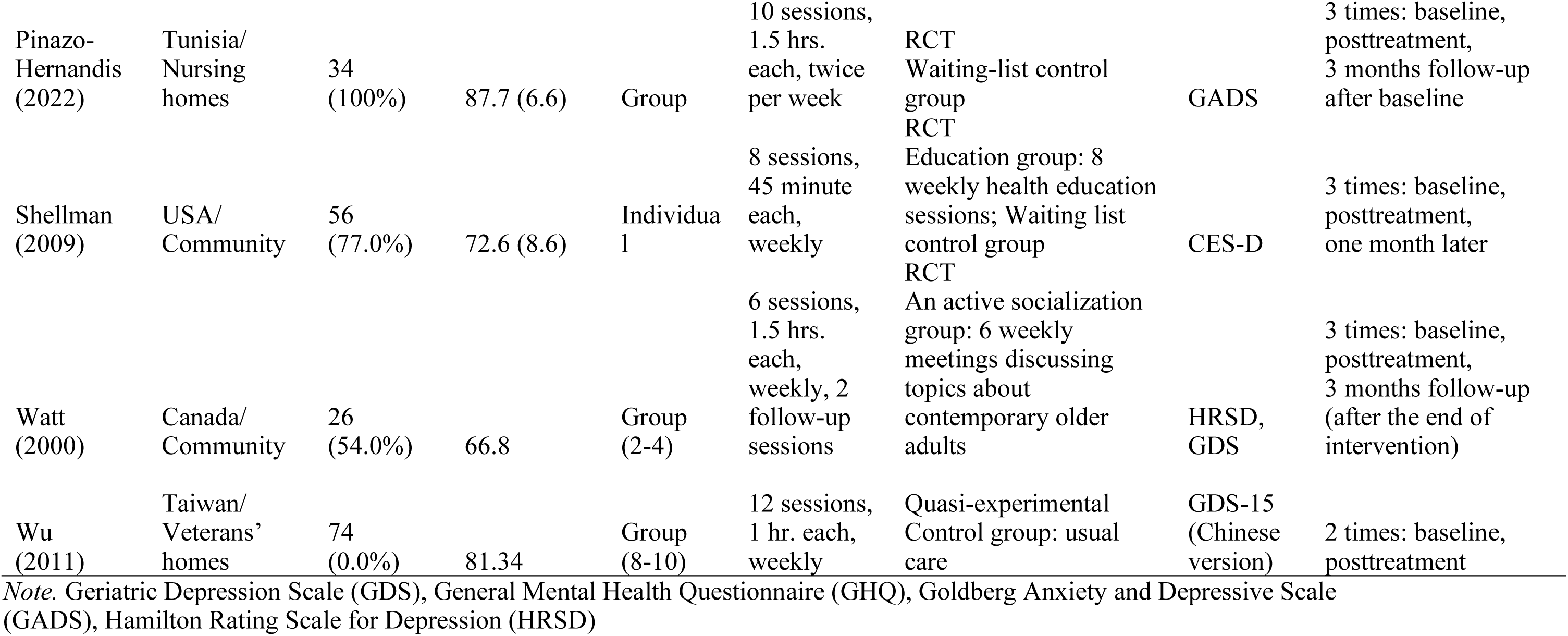
Study Characteristics (N=9)

### Risk of Bias in Studies

All six randomized controlled trials were assessed using the ROB 2 tool. While all studies employed random assignment of participants, however, the methods for allocation concealment were not clearly reported. In four studies (Pearson, 2006; Pinazo-Hernandis et al., 2022; Sabir et al., 2016; Shellman et al., 2009), outcome assessors were aware of the intervention received by participants, which could potentially influence the assessment of outcomes. Besides the randomization process and outcome measurement, more than 66.7% of the included studies were rated as low risk of bias in all the other domains. Figure 2 outlines the quality appraisal results for RCTs.

**Figure 2.**
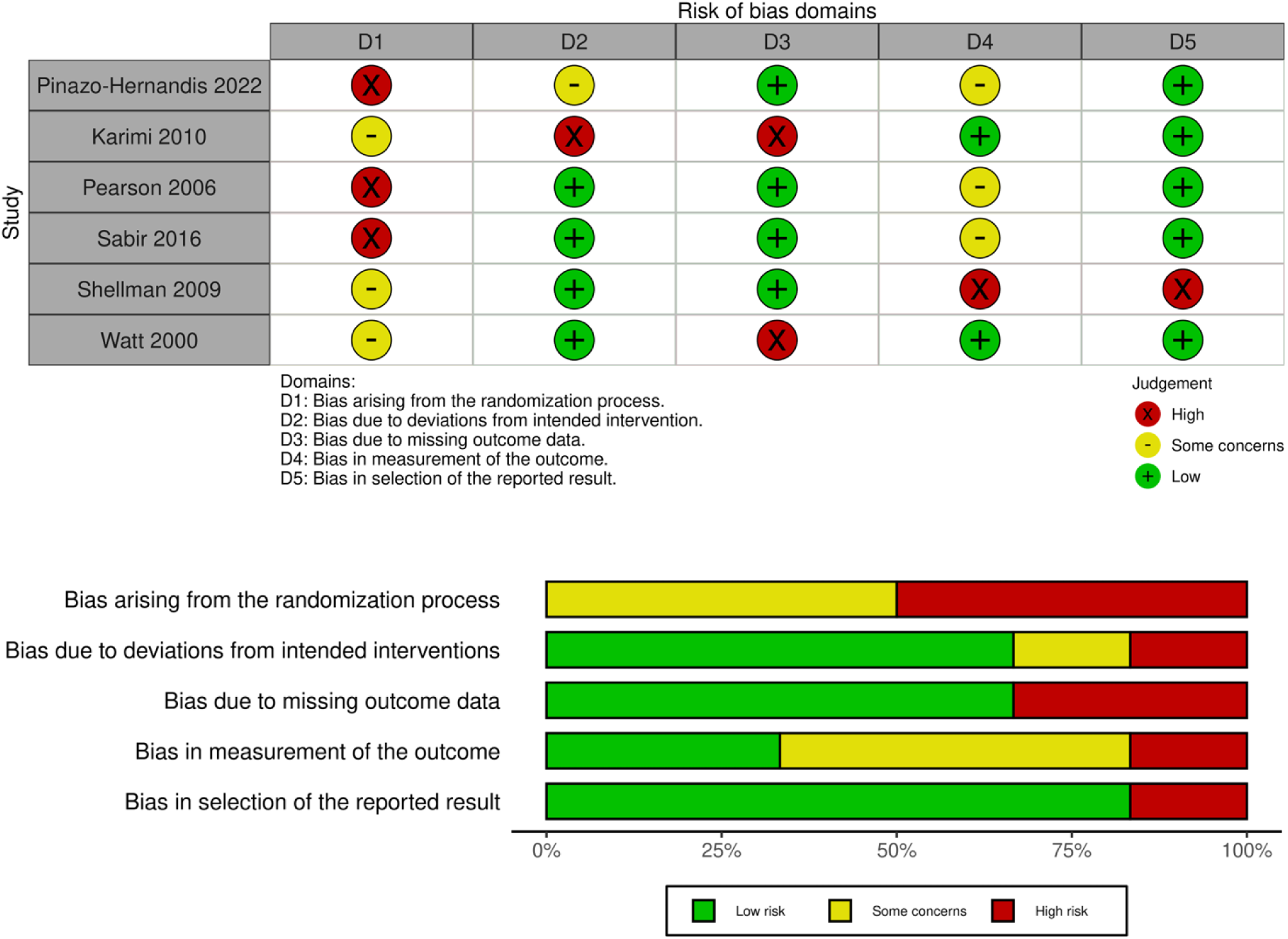
Risk of Bias in RCTs by ROB 2 tool (n = 6)

The ROBINS-I tool was used to assess the risk of bias in five quasi-experimental studies, all employing two-arm designs with control groups. Although confounding was a common concern due to the non-randomized nature of these studies, Bohlmeijer et al. (2009), Musavi et al. (2017), and Wu (2011) applied block randomization, stratification, and regression models to mitigate bias. The moderate risk of bias rating for outcome measure reflects the standard characteristics of studies utilizing participant-reported scales and does not imply any deficiency in their quality. Over 75% of the studies were rated as low risk of bias across the remaining five domains. Overall, all studies were rated as low risk of bias, with quality appraisal results summarized in Figure 3.

**Figure 3.**
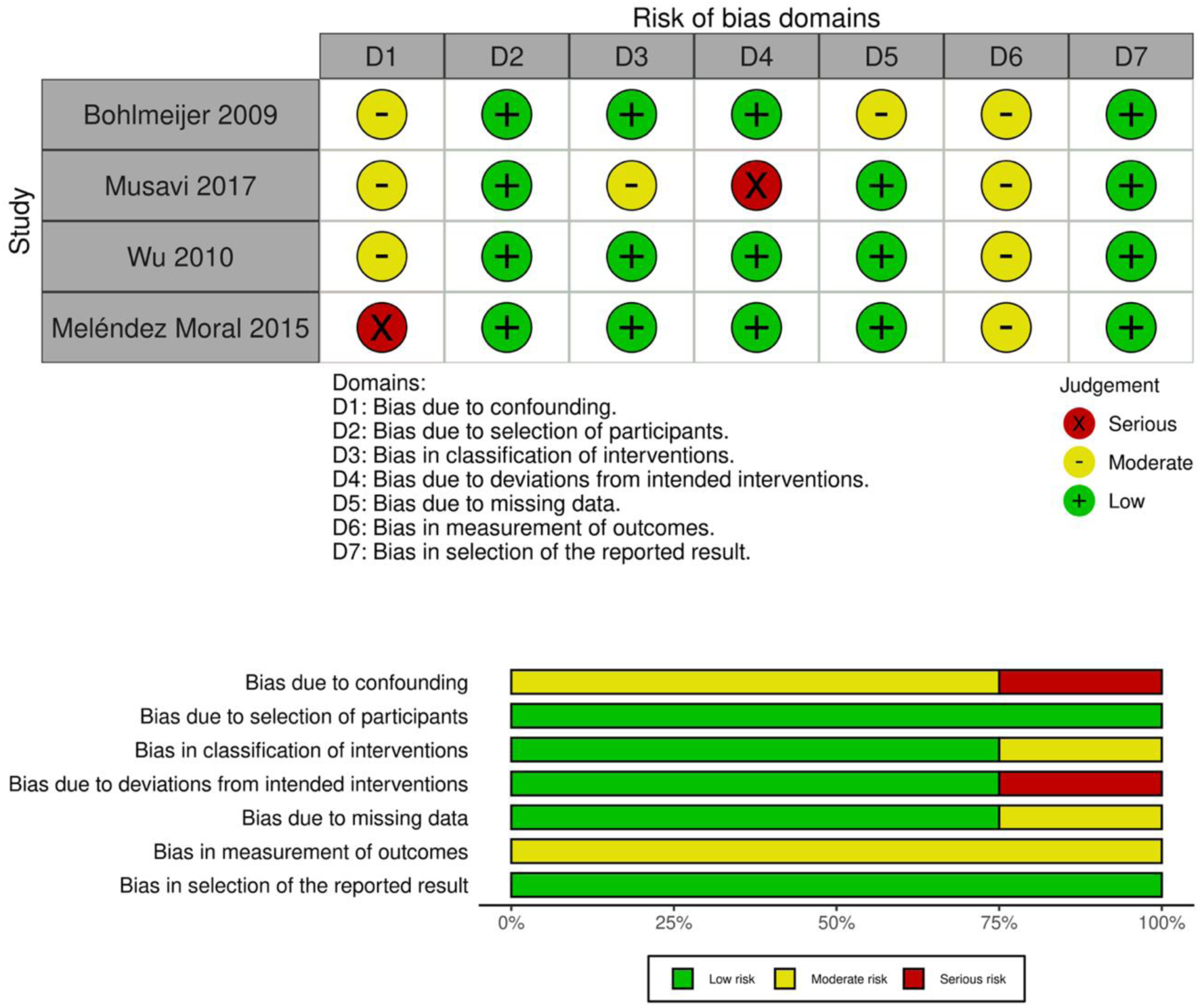
Risk of Bias in Quasi-experimental studies by ROBINS-I tool (n = 4)

### Results of Syntheses

#### The Effect of Integrative Reminiscence

Among the nine articles included in this meta-analysis, four demonstrate statistically significant differences in the effect of integrative reminiscence compared to the corresponding control in reducing depression from post-baseline to baseline. The effect sizes across these studies ranged from 0.28 to 1.62 (see Figure 4).

**Figure 4.**
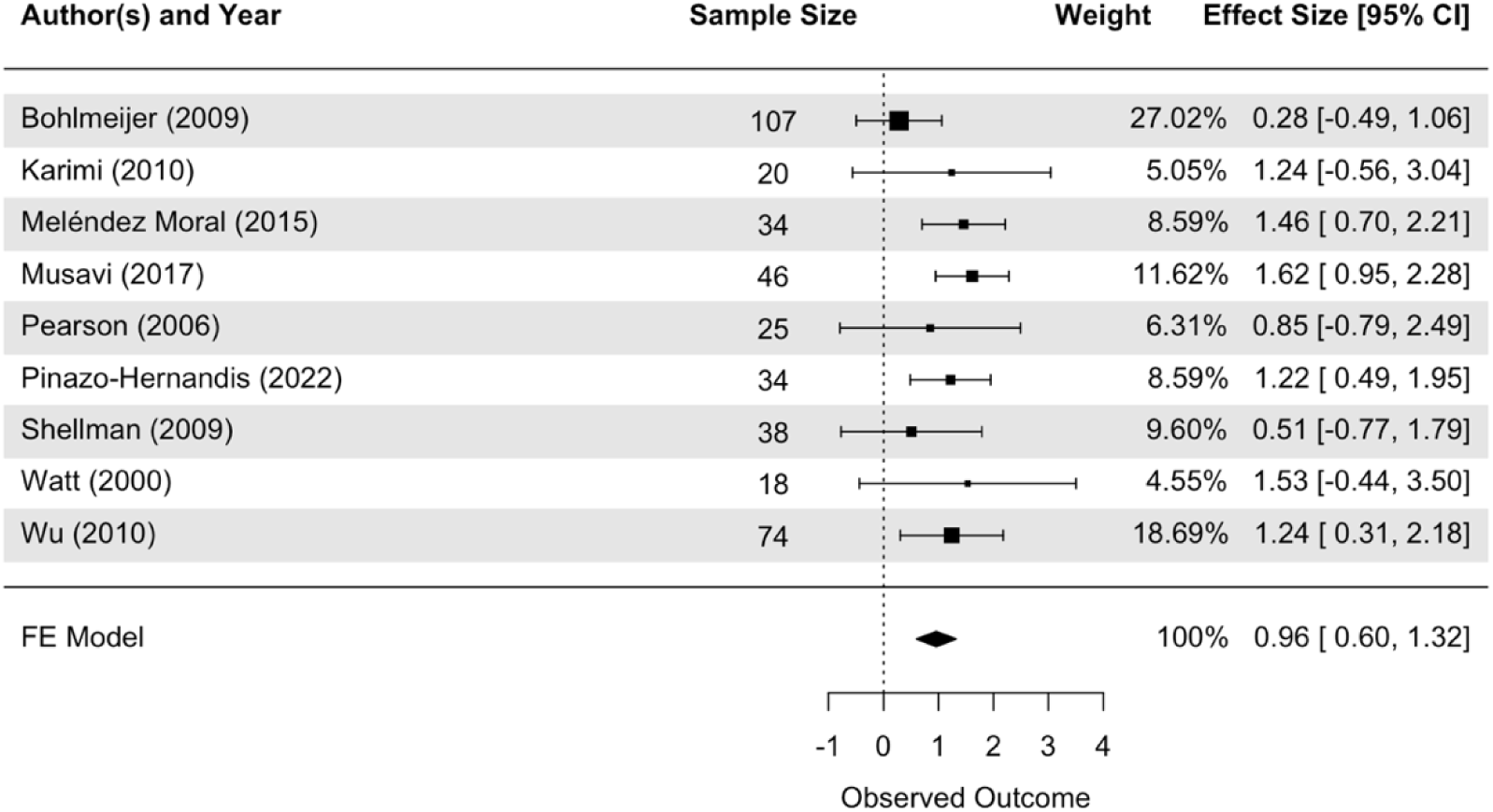
Forest Plot of the Effects of Integrative Reminiscence Interventions on Depressive Symptoms.

A fixed-effects model is chosen considering insufficient evidence implying heterogeneity (Q = 8.6219; I^2 = 7.21%; p = .3752). The fixed-effects model shows that integrative reminiscence has a larger effect on depression reduction compared to control (Cohen’s d = 0.9608; z = 5.2049; p < .0001), with a 95% confidence interval between 0.5990 and 1.3225 and a standard error of 0.1846, which implies a large overall effect.

### Subgroup Analysis

#### Intervention Group Size

Figure 5a shows the results of subgroup analysis. For the subgroup analysis, the integrative reminiscence in large group form (total sample size = 140) exhibits significant effects in reducing depression (Cohen’s d = 1.3644; 95% CI: (0.7651, 1.9637); p < .0001), while the reminiscence in small group form (total sample size = 163) results in statistically non-significant effect in depression reduction (Cohen’s d = 0.4738; 95% CI: (-0.1554, 1.1030); p = .1400). For the Unknown group (total sample size = 93), the combined effect is also statistically significant (Cohen’s d = 1.2066; 95% CI: (0.6209, 1.7923); p < .0001). The effect size for the integrative reminiscence in large group form is significantly larger than those in small group form (difference in Cohen’s d = 0.8906; 95% CI: (0.0217, 1.7596); p = .0445). In addition, no heterogeneity is detected in any subgroups with the p values of .7888, .5109, and .7781 for large, small group, and unknown groups.

**Figure 5a.**
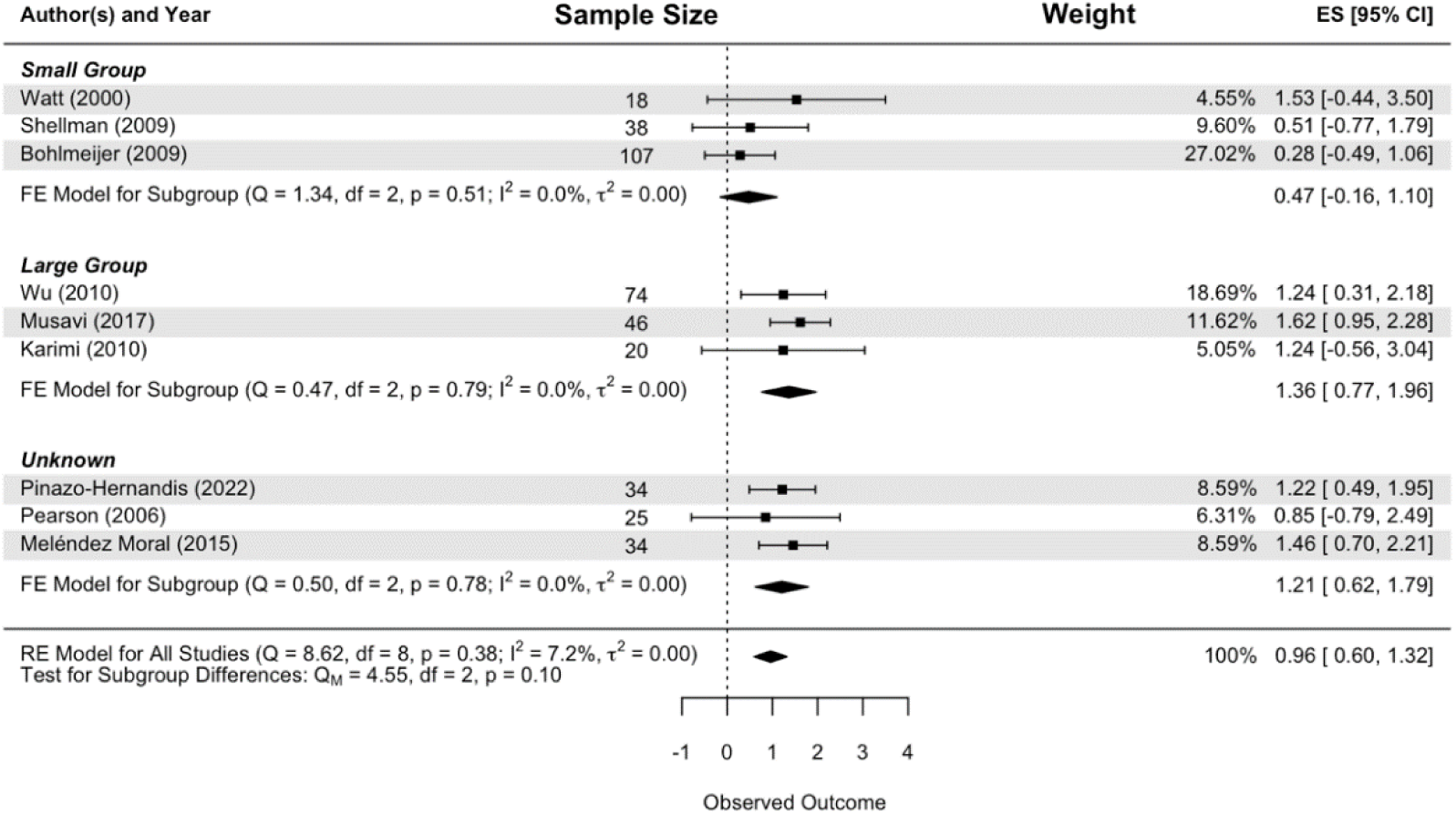
Forest Plot of the Subgroup Analysis for Intervention Group Size.

#### The Setting of the Intervention

Statistical significance for the effectiveness of the integrative reminiscence has been observed in both community settings (total sample size = 222, Cohen’s d = 0.6668; 95% CI: (0.1559, 1.1777); p = .0105) and nursing home setting (total sample size = 174, Cohen’s d = 1.3358; 95% CI: (0.8328, 1.8388); p < .0001). The effect size for the integrative reminiscence in the nursing home settings is larger than those in community settings (difference in Cohen’s d = 0.6690; 95% CI: (0.1172, 1.4552); p = .0953), although the differential effect does not reach significance. No heterogeneity is detected for either intervention setting (See Figure 5b).

**Figure 5b.**
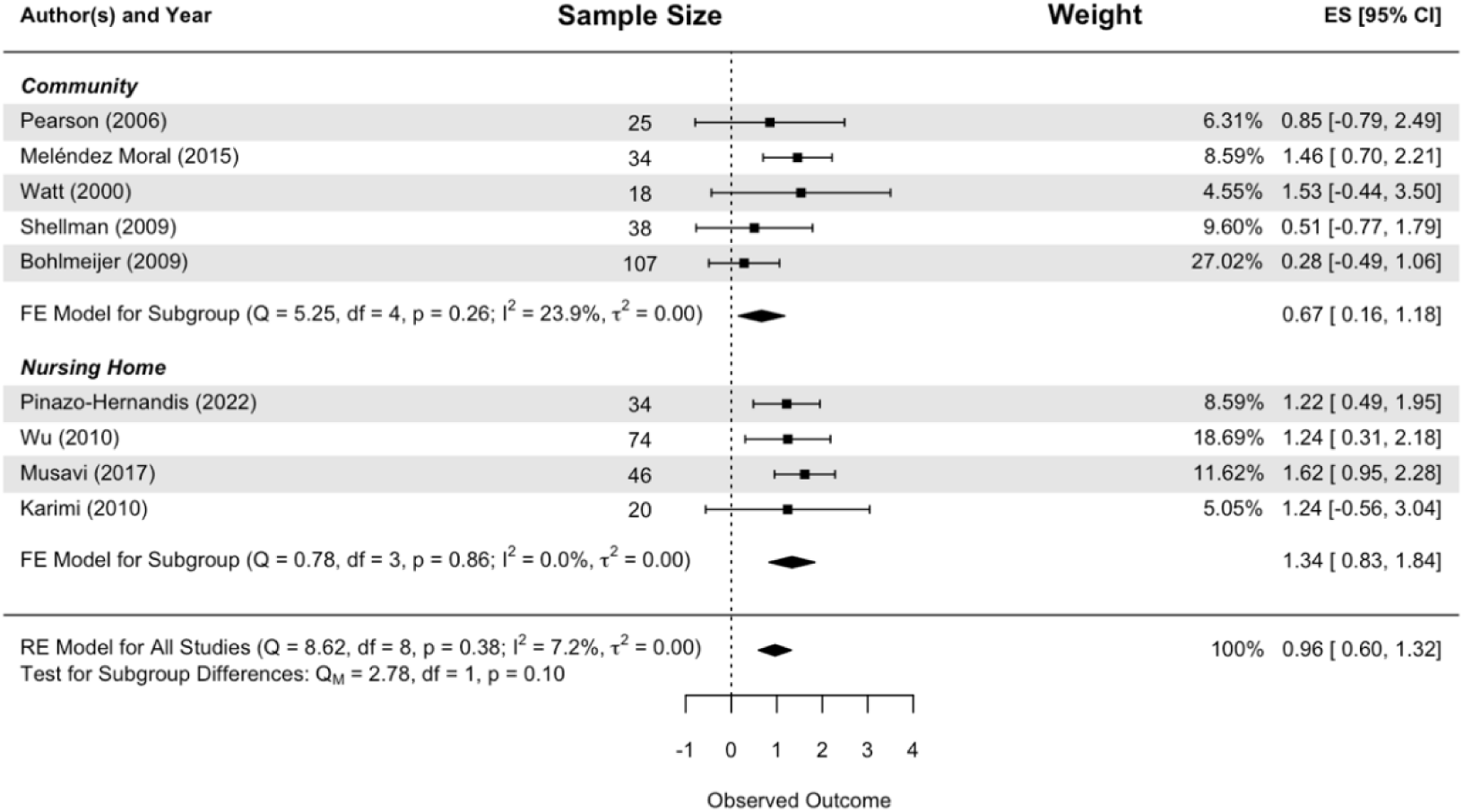
Forest Plot of the Subgroup Analysis for the Setting of the Intervention.

#### Type of Control Group

Both the active control subgroup (total sample size = 76, Cohen’s d = 0.9436; 95% CI: (0.0206, 1.8666); p = .0451) and waiting-list control subgroup (total sample size = 320, Cohen’s d = 0.9648; 95% CI: (0.5745, 1.3552); p < .0001) produce similar effect sizes as the overall effect. Their Cohen’s d only differs by 0.0212 with a p-value of .9699 and 95% CI: (-1.0800, 1.1224). No heterogeneity is detected for either type of control group. Figure 5c illustrates the subgroup analysis for different types of control groups.

**Figure 5c.**
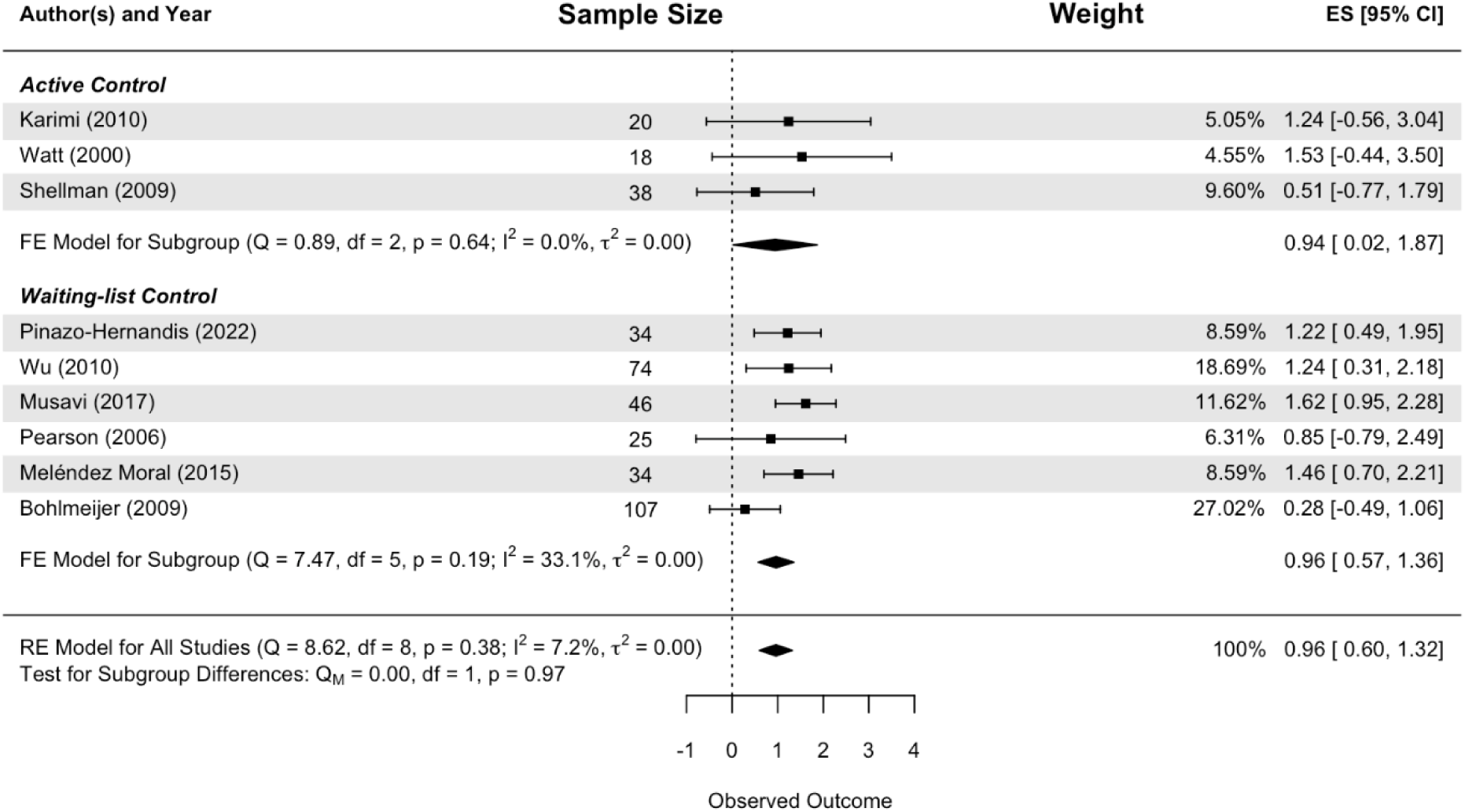
Forest Plot of the Subgroup Analysis for Type of Control Group.

### Publication Bias

A funnel plot is depicted in Figure 6, demonstrating a low risk of publication bias through visual inspection. Egger’s regression test for the funnel plot asymmetry does not yield a significant result (*z* = 0.5598, *p* = .5756).

**Figure 6.**
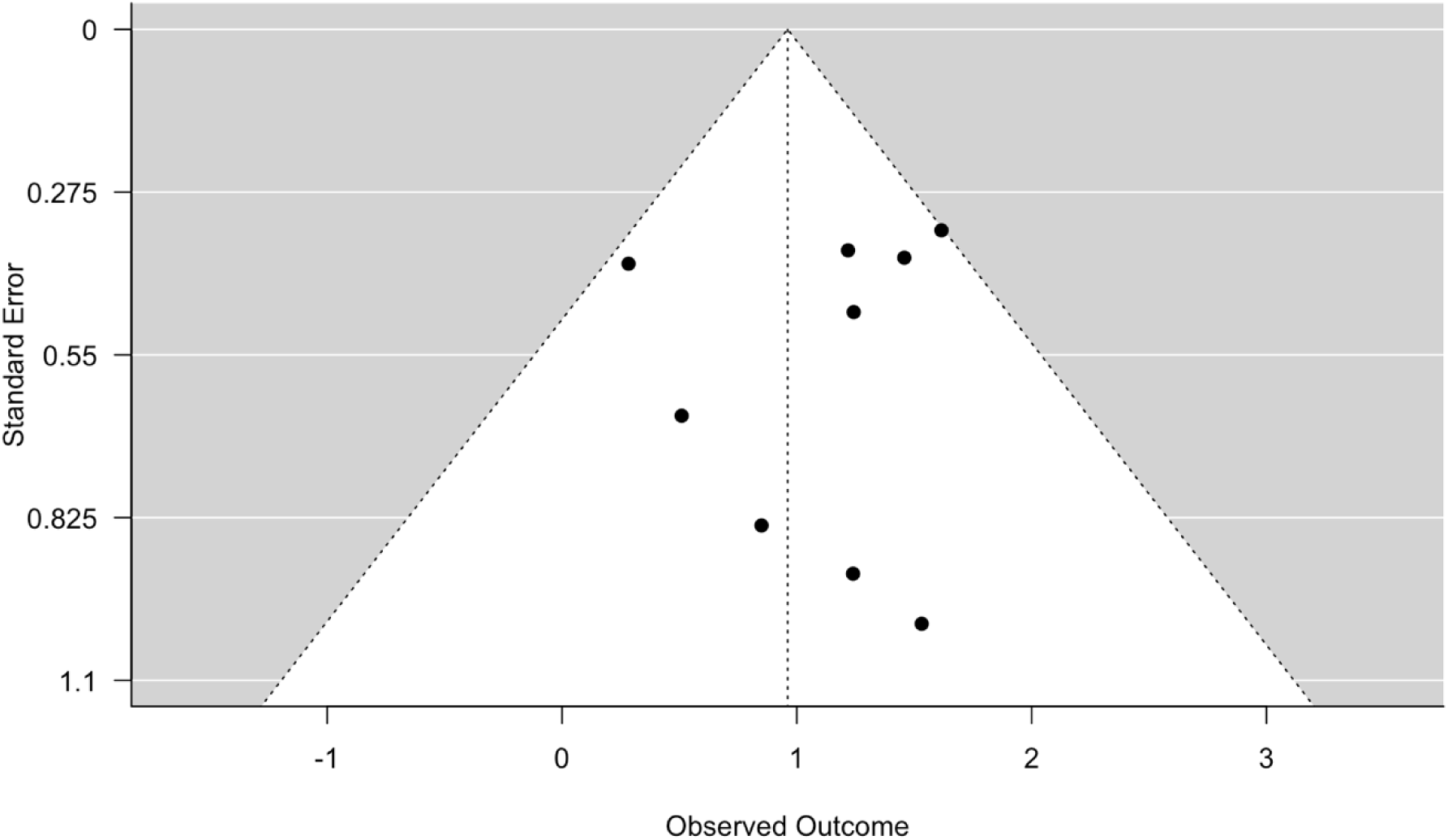
Publication Bias Assessment by Funnel Plot.

## Discussion

The current review synthesized the results from nine studies involving 396 older adults. The meta-analysis showed a large and statistically significant effect of integrative reminiscence in reducing depression from post-baseline to baseline (Cohen’s *d* = 0.9608; *p* < .0001). The effects at 3-month follow-up or longer showed a similarly large effect size (Cohen’s *d* = 1.3159; *p* < .0001), indicating the effect of integrative reminiscence could be sustained over time for older adults with depressive symptoms. The significant and large effects of integrative reminiscence intervention are much larger than the size of the effect reported by Tam et al. (2021, Cohen’s *d* = -0.38; *p* < .0001) in their meta-analysis, which included a variety of reminiscence interventions, showing integrative reminiscence is salient in reminiscence interventions in depression reduction.

The effect sizes examined for integrative reminiscence are comparable to those found in well-established treatments, such as cognitive behavioral therapy. For example, Gould et al. (2012) found a large and statistically significant effect size of -1.35 when cognitive behavioral therapy was compared to non-active controls. However, when compared to active controls, the cognitive behavioral therapy was not significantly more effective in reducing depressive symptoms (Gould et al., 2012). In the current review, significant results have been exhibited when comparing the integrative reminiscence intervention to active controls, which indicates integrative reminiscence may have more substantial evidence of effectiveness compared to cognitive behavioral therapy. This finding suggests that integrative reminiscence reduces depressive symptomatology not only through nonspecific therapeutic factors as other psychotherapy (e.g., attention or empathy received from facilitators, the human interaction, or social support between group members) (Mohr et al., 2009) but also through specific elements of integrative reminiscence.

In analyzing subgroups, we examined the impact of group size and found that the effect size for integrative reminiscence when conducted in a larger group (n > 4) exceeds that observed in smaller group contexts, and there presents a statistically significant (*p* = .0445) differential effect that integrative reminiscence is more effective when conducted in a larger group setting. A larger group technique can better utilize the advantage of a group format and exert its social functions. The presence of more individuals in a group can provide more opportunities for social interaction and engagement (Hanaoka et al., 2011). Increased interactions can be beneficial in reducing feelings of loneliness and isolation and promoting a sense of belonging and community among the participants. A larger group increases the chances for older adults to receive feedback and affirmation from peers, maximizing the benefits of integrative reminiscence intervention.

Though not statistically significant at a 5% significance level (p-value = .0953), a medium to large effect size of 0.6690 indicates that integrative reminiscence has a larger effect on alleviating depressive symptoms among nursing home residents compared to older adults living in the community. This finding contrasts with the conclusion suggested by Bohlmeijer et al. (2007) that reminiscence interventions are more effective for older adults living in the community. Bohlmeijer et al.’s (2007) analysis investigated the combined effect of the various reminiscence interventions, which mainly included non-specific reminiscence. Compared to non-specific reminiscence or simple reminiscence programs, integrative reminiscence contains construction healing components and is thus more powerful. Therefore, integrative reminiscence is more suitable for helping distressed old adults, such as those with depression, to help them accept negative past events and integrate a continuity between the past and present. Considering the relatively worse mental health in nursing home residents, it is reasonable that integrative reminiscence showed a greater effect on older adults living in nursing homes. Future research is needed to confirm these findings.

In the earlier meta-analysis studies that examined the effect of an assorted collection of the various types of reminiscence-based intervention, high heterogeneity remained a problem (Bohlmeijer et al., 2003; Liu et al., 2021; Tam et al., 2021; Westerhof & Slatman, 2019). High heterogeneity limits the interpretability and usability of even rigorously conducted studies (Imrey, 2020). Unlike the earlier meta-analyses, the heterogeneity was low (I^2 = 30.39%;) and non-significant (*Q* = 11.4926; *p* = .1753) in this meta-analysis. These findings provide additional substantiation for our assertion that incorporating all reminiscence interventions introduces statistical heterogeneity. Examining the efficacy of reminiscence on depression in older adults without accounting for the diversity in reminiscence interventions renders the conclusions on the efficacy of reminiscence inconclusive and less convincing. Hence, we emphasize the importance of differentiating the various types of reminiscence in research and clinical practice.

Integrative reminiscence was examined as an effective treatment for depressive symptoms in older adults, offering a crucial alternative to conventional psychotherapies. Thus, integrative reminiscence should be adopted as a standard form of psychotherapy. Due to its high adaptability to various mental health care settings, it can serve as a part of routine care for institutionalized elderly and as a community-based intervention in community centers (Bohlmeijer et al., 2007; Watt & Cappeliez, 2000). Moreover, integrative reminiscence can be applied as a standard therapy in psychiatric clinics to treat depression. Integrative reminiscence has the merit to aid in alleviating psychological distress in hospice or palliative care settings. Further research is needed to evaluate the effectiveness of the intervention in hospice or palliative care settings.

## Limitations

The scope of the literature search was limited to published articles in English. Unpublished articles and studies published in other languages were not searched. There is a possibility that some articles on this topic may be omitted. The inclusion of unpublished data may have altered effect sizes and, hence, conclusions about the efficacy of the intervention, although there was no evidence of publication bias in the current study.

The risk of bias, assessed via ROB 2 for RCTs and ROBINS-I for non-RCTs, identified concerns across several domains. Notably, the RCT studies lacked detailed randomization processes and exhibited potential biases due to limitations on trial compliance and the presence of missing data. Outcome measurement was compromised by unblinded assessors in some cases, and selective result reporting was evident. Non-RCTs varied in confounding risk, with some studies not adequately controlling for potential confounders and others presenting unclear intervention classifications and deviations, alongside issues with missing data handling. While the studies generally met acceptable quality standards, the concerns identified are non-trivial. These findings highlight the need for a more rigorous and stringent framework to standardize clinical trials and reporting to reduce bias and enhance result reliability. In the data synthesis stage, the weights assigned to each study only depend on the sample sizes of the trials and were not adjusted according to the quality of the studies, which could potentially underestimate the variation when making inferences. Nevertheless, the meta-analysis results are consistent, and analyses indicate no heterogeneity or evidence of publication bias.

## Conclusion

This review is important in that it is the first review that focuses on the effect of integrative reminiscence on depression in older adults. The results demonstrate the significantly large effect of this intervention in alleviating depressive symptoms and provide solid empirical evidence for employing integrative reminiscence to reduce depressive symptoms in older adults. This review concluded that integrative reminiscence is an effective treatment for depressive symptoms in older adults and holds the potential to serve as a substitute for other forms of psychotherapy.

## Data Availability

All data produced in the present work are contained in the manuscript.

## Registration and Protocol

The study protocol is registered in the International Prospective Register of Systematic Reviews (ID: CRD42023452477).

## Ethics Statement

Ethical approval was not required for this study, as it is a systematic review based only on publicly available data from published research, with no direct involvement of human participants.

## Acknowledgments

None.

## References

American Psychiatric Association. (2013). Diagnostic and statistical manual of mental disorders (5th ed.).

Aşiret, G. D., & Kapucu, S. (2016). The effect of reminiscence therapy on cognition, depression, and activities of daily living for patients with Alzheimer disease. Journal of Geriatric Psychiatry and Neurology, 29(1), 31–37. 10.1177/0891988715598233

Baquero, M. (2015). Depressive symptoms in neurodegenerative diseases. World Journal of Clinical Cases, 3(8), 682. 10.12998/wjcc.v3.i8.682

Bohlmeijer, E. T., Kramer, J., Smit, F., Onrust, S., & H, van M. (2009). The effects of integrative reminiscence on depressive symptomatology and mastery of older adults. Community Mental Health Journal, 45(6), 476–484. 10.1007/s10597-009-9246-z

Bohlmeijer, E. T., Roemer, M., Cuijpers, P., & Smit, F. (2007). The effects of reminiscence on psychological well-being in older adults: A meta-analysis. In Aging and Mental Health (Vol. 11, Issue 3). 10.1080/13607860600963547

Bohlmeijer, E. T., Smit, F., & Cuijpers, P. (2003). Effects of reminiscence and life review on late-life depression: A meta-analysis. International Journal of Geriatric Psychiatry, 18(12), 1088–1094. 10.1002/gps.1018

Bohlmeijer, E. T., Valenkamp, M., Westerhof, G., Smit, F., & Cuijpers, P. (2005). Creative reminiscence as an early intervention for depression: Results of a pilot project. Aging and Mental Health, 9(4), 302–304. 10.1080/13607860500089567

Bohlmeijer, E. T., Westerhof, G. J., & Emmerik-de Jong, M. (2008). The effects of integrative reminiscence on meaning in life: Results of a quasi-experimental study. Aging & Mental Health, 12(5), 639–646. 10.1080/13607860802343209

Butler, R. N. (1963). The Life Review: An Interpretation of Reminiscence in the Aged. Psychiatry, 26(1), 65–76. 10.1080/00332747.1963.11023339

Gonzalez, J., Mayordomo, T., Torres, M., Sales, A., & Meléndez, J. C. (2015). Reminiscence and dementia: A therapeutic intervention. International Psychogeriatrics, 27(10), 1731– 1737. 10.1017/S1041610215000344

Gould, R. L., Coulson, M. C., & Howard, R. J. (2012). Cognitive behavioral therapy for depression in older people: A meta-analysis and meta-regression of randomized controlled trials. Journal of the American Geriatrics Society, 60(10), 1817–1830. 10.1111/j.1532-5415.2012.04166.x

Haidich, A. B. (2014). Meta-analysis in medical research Meta-analysis in medical research. Hippokratia, 14(1), 29–37. http://scholar.google.com

Hanaoka, H., Muraki, T., Yamane, S., Shimizu, H., & Okamura, H. (2011). Testing the feasibility of using odors in reminiscence therapy in Japan. Physical and Occupational Therapy in Geriatrics, 29(4), 287–299. 10.3109/02703181.2011.628064

Higgins, J. P., Savović, J., Page, M. J., & Sterne, J. A. C. (2019). The Cochrane Collaboration’s tool for assessing risk of bias in randomised trials. British Medical Journal, *July*, 1–24. https://methods.cochrane.org/

Hofer, J., Busch, H., Poláčková Šolcová, I., & Tavel, P. (2017). When Reminiscence is Harmful: The Relationship Between Self-Negative Reminiscence Functions, Need Satisfaction, and Depressive Symptoms Among Elderly People from Cameroon, the Czech Republic, and Germany. Journal of Happiness Studies, 18(2), 389–407. 10.1007/s10902-016-9731-3

Huang, H. C., Chen, Y. T., Chen, P. Y., Huey-Lan Hu, S., Liu, F., Kuo, Y. L., & Chiu, H. Y. (2015). Reminiscence Therapy Improves Cognitive Functions and Reduces Depressive Symptoms in Elderly People With Dementia: A Meta-Analysis of Randomized Controlled Trials. Journal of the American Medical Directors Association, 16(12), 1087–1094. 10.1016/j.jamda.2015.07.010

Imrey, P. B. (2020). Limitations of Meta-analyses of Studies with High Heterogeneity. JAMA Network Open, 3(1), 2019–2021. 10.1001/jamanetworkopen.2019.19325

Irazoki, E., García-Casal, J. A., Sánchez Meca, J., & Franco Martín, M. (2017). Efficacy of group reminiscence therapy for people with dementia. Systematic literature review and meta-analysis. Rev Neurol, 65(10), 447–456. 10.33588/rn.6510.2017381

Karimi, H., Dolatshahee, B., Momeni, K., Khodabakhshi, A., Rezaei, M., & AA, K. (2010). Effectiveness of integrative and instrumental reminiscence therapies on depression symptoms reduction in institutionalized older adults: An empirical study. Aging & Mental Health, 14(7), 881–887. 10.1080/13607861003801037

Kovach, C. (1991). Reminiscence: Exploring the Origins, Processes, and Consequences. Nursing Forum, 26(3), 14–20. 10.1111/j.1744-6198.1991.tb00884.x

Lieberman, M. A., & Tobin, S. S. (1983). The experience of old age: stress, coping and survival. Basic Books.

Liu, Z., Yang, F., Lou, Y., Zhou, W., & Tong, F. (2021). The Effectiveness of Reminiscence Therapy on Alleviating Depressive Symptoms in Older Adults: A Systematic Review. Frontiers in Psychology, 12. 10.3389/fpsyg.2021.709853

Lüdecke, D. (2018). esc: Effect Size Computation for Meta Analysis (0.4.1). Zenodo. 10.5281/zenodo.1249218

McGuinness, L. A., & Higgins, J. P. T. (2021). Risk-of-bias VISualization (robvis): An R package and Shiny web app for visualizing risk-of-bias assessments. Research Synthesis Methods, 12(1), 55–61. 10.1002/jrsm.1411

Meléndez Moral, J. C., Fortuna Terrero, F. B., Sales Galán, A., & Mayordomo Rodríguez, T. (2015). Effect of integrative reminiscence therapy on depression, well-being, integrity, self-esteem, and life satisfaction in older adults. The Journal of Positive Psychology, 10(3), 240–247. 10.1080/17439760.2014.936968

Meyer, C., & O’Keefe, F. (2020). Non-pharmacological interventions for people with dementia: A review of reviews. Dementia, 19(6), 1927–1954. 10.1177/1471301218813234

Mohr, D. C., Spring, B., Freedland, K. E., Beckner, V., Arean, P., Hollon, S. D., Ockene, J., & Kaplan, R. (2009). The selection and design of control conditions for randomized controlled trials of psychological interventions. Psychotherapy and Psychosomatics, 78(5), 275–284. 10.1159/000228248

Musavi, M., Mohammadian, S., & Mohammadinezhad, B. (2017). The effect of group integrative reminiscence therapy on mental health among older women living in Iranian nursing homes. Nursing Open, 4(4), 303–309. 10.1002/nop2.101

Nina, R. E. (2014). P2-143: Depression and Vascular Dementia. Alzheimer’s & Dementia, 10(4S_Part_13), 2–3. 10.1016/j.jalz.2014.05.818

O’ Philbin, L., Woods, B., Farrell, E. M., Spector, A. E., & Orrell, M. (2018). Reminiscence therapy for dementia: an abridged Cochrane systematic review of the evidence from randomized controlled trials. Expert Review of Neurotherapeutics, 18(9), 715–727. 10.1080/14737175.2018.1509709

Oluboka, O. J., Katzman, M. A., Habert, J., McIntosh, D., MacQueen, G. M., Milev, R. V., McIntyre, R. S., & Blier, P. (2018). Functional recovery in major depressive disorder: Providing early optimal treatment for the individual patient. International Journal of Neuropsychopharmacology, 21(2), 128–144. 10.1093/ijnp/pyx081

Parker, R. G. (1995). Reminiscence : A Continuity Theory Framework. 35(4), 515–525.

Pearson, L. (2006). The effect of integrative reminiscence on depression, ego integrity and personal mastery in the elderly. In ProQuest Dissertations Publishing.

Petersen, J. D., Waldorff, F. B., Siersma, V. D., Phung, T. K. T., Bebe, A. C. K. M., & Waldemar, G. (2017). Major depressive symptoms increase 3-year mortality rate in patients with mild dementia. International Journal of Alzheimer’s Disease, 2017. 10.1155/2017/7482094

Pinazo-Hernandis, S., Sales, A., & Martinez, D. (2022). Older Women’s Loneliness and Depression Decreased by a Reminiscence Program in Times of COVID-19. Frontiers in Psychology, 13(February), 1–9. 10.3389/fpsyg.2022.802925

R Core Team. (2023). R: A language and environment for statistical computing. R Foundation for Statistical Computing.

Sabir, M., Henderson, C. R., Kang, S.-Y., & Pillemer, K. (2016). Attachment-focused integrative reminiscence with older African Americans: a randomized controlled intervention study. Aging & Mental Health, 20(5), 517–528. 10.1080/13607863.2015.1023764

Shellman, J. M. (2016). Examining Patterns and Functions of Reminiscence in a Sample of Black Adults: Implications for Psychiatric Nurses. Archives of Psychiatric Nursing, 30(3), 387–392. 10.1016/j.apnu.2016.01.006

Shellman, J. M., Mokel, M., & Hewitt, N. (2009). The effects of integrative reminiscence on depressive symptoms in older African Americans. Western Journal of Nursing Research, 31(6), 772–786. 10.1177/0193945909335863

Sterne, J. A., Hernán, M. A., Reeves, B. C., Savović, J., Berkman, N. D., Viswanathan, M., Henry, D., Altman, D. G., Ansari, M. T., Boutron, I., Carpenter, J. R., Chan, A. W., Churchill, R., Deeks, J. J., Hróbjartsson, A., Kirkham, J., Jüni, P., Loke, Y. K., Pigott, T. D., … Higgins, J. P. (2016). ROBINS-I: A tool for assessing risk of bias in non-randomised studies of interventions. BMJ (Online), 355, 4–10. 10.1136/bmj.i4919

Tam, W., Poon, S. N., Mahendran, R., Kua, E. H., & Wu, X. V. (2021). The effectiveness of reminiscence-based intervention on improving psychological well-being in cognitively intact older adults: A systematic review and meta-analysis. International Journal of Nursing Studies, 114, 103847. 10.1016/j.ijnurstu.2020.103847

Van Bogaert, P., Tolson, D., Eerlingen, R., Carvers, D., Wouters, K., Paque, K., Timmermans, O., Dilles, T., & Engelborghs, S. (2016). SolCos model-based individual reminiscence for older adults with mild to moderate dementia in nursing homes: a randomized controlled intervention study. Journal of Psychiatric & Mental Health Nursing (John Wiley & Sons, Inc.), 23(9/10), 568–575. 10.1111/jpm.12336

Viechtbauer, W. (2010). Conducting Meta-Analyses in R with the metafor Package. Journal of Statistical Software, 36(3), 1–48. 10.18637/jss.v036.i03

Watt, L. M., & Cappeliez, P. (1995). Reminiscence interventions for the treatment of depression in older adults. In The art and science of reminiscing: Theory, research, methods, and applications. (pp. 221–232). Taylor & Francis.

Watt, L. M., & Cappeliez, P. (2000). Integrative and instrumental reminiscence therapies for depression in older adults: Intervention strategies and treatment effectiveness. Aging and Mental Health, 4(2), 166–177. 10.1080/13607860050008691

Watt, L. M., & Wong, P. T. P. (1991). A taxonomy of reminiscence and therapeutic implications. Journal of Gerontological Social Work, 16(1–2), 37–57. 10.1300/J083v16n01_04

Webster, J. D., Bohlmeijer, E. T., & Westerhof, G. J. (2010). Mapping the future of reminiscence: A conceptual guide for research and practice. Research on Aging, 32(4), 527–564. 10.1177/0164027510364122

Westerhof, G. J., & Slatman, S. (2019). In search of the best evidence for life review therapy to reduce depressive symptoms in older adults: A meta-analysis of randomized controlled trials. Clinical Psychology: Science and Practice, 26(4). 10.1111/cpsp.12301

Woods, B., O’Philbin, L., Farrell, E. M., Spector, A. E., & Orrell, M. (2018). Reminiscence therapy for dementia. Cochrane Database of Systematic Reviews. 10.1002/14651858.CD001120.pub3

World Health Organization (WHO). (2022). Ageing and health. World Health Organization (WHO). https://www.who.int/news-room/fact-sheets/detail/ageing-and-health

Wu, L.-F. (2011). Group integrative reminiscence therapy on self-esteem, life satisfaction and depressive symptoms in institutionalised older veterans. Journal of Clinical Nursing (John Wiley & Sons, Inc.), 20(15–16), 2195–2203. 10.1111/j.1365-2702.2011.03699.x

Yang, Youngsoon. (2017). [P3–443]: The Relationship Between the Geriatric Depression Scale and Cognitive-Behavioral Aspects in ALZHEIMER’s DEMENTIA. Alzheimer’s & Dementia, 13(7S_Part_23), 2017. 10.1016/j.jalz.2017.06.1661

Yang, Yuxuan, Graf, L., Longdin, M., Khait, A. A., & Shellman, J. (2022). Scoping review of reminiscence research undertaken in long-term care communities. Geriatric Nursing, 46, 191–198. 10.1016/j.gerinurse.2022.06.004

