## Appendix A for "The Effect of Integrative Reminiscence in Older Adults with Depression: A Systematic Review and Meta-Analysis"

**Appendix A. Search Methods**

**PubMed**

Search: ((Aged[Mesh] OR elderly[Text Word] OR aged[Text Word] OR older[Text Word] OR elder[Text Word] OR geriatric[Text Word] OR elderly people[Text Word] OR old people[Text Word] OR older people[Text Word] OR senior[Text Word]) AND ("Depression"[Mesh] OR depression[Title/Abstract] OR depressed[Title/Abstract] OR depressive[Title/Abstract])) AND (Integrative reminiscence therapy[Title/Abstract] OR Integrative reminiscence[Title/Abstract] OR Integrative reminisce[Title/Abstract] OR life review[Title/Abstract] OR life story[Title/Abstract]) Filters: English, prior to 20231231

Aged[Mesh]: "aged"[MeSH Terms]

**CINAHL Plus with Full Text**
((MH "Depression+") OR (MH "Geriatric Depression Scale") OR (MH "Center for Epidemiological Studies Depression Scale") ) OR ( AB depression or depressive disorder or depressive symptoms or major depressive disorder) ) AND ( (TI Integrative Reminiscence) OR (AB Integrative Reminiscence) OR (MH Reminiscence Therapy (Iowa NIC)) OR (MH Reminiscence Therapy) OR (MJ life review) OR (AB life story) OR (AB life review) ) AND ( MH "Aged+") OR (MH "Gerontologic Nursing+") OR (MH "Gerontologic Care") OR TX ( aged or elderly or senior or older people or geriatric or gerontologic or older adults) )

**Limiters** - Published Date: prior to 20231231

**Expanders** - Apply equivalent subjects

**Narrow by Language:** - English

**Search modes** - Boolean/Phrase

**PsycInfo**

((MH "Depression+") OR (MH "Geriatric Depression Scale") OR (MH "Center for Epidemiological Studies Depression Scale") ) OR ( AB depression or depressive disorder or depressive symptoms or major depressive disorder) ) AND ( (TI Integrative Reminiscence) OR (AB Integrative Reminiscence) OR (MH Reminiscence Therapy (Iowa NIC)) OR (MH Reminiscence Therapy) OR (MJ life review) OR (AB life story) OR (AB life review) ) AND ( MH "Aged+") OR (MH "Gerontologic Nursing+") OR (MH "Gerontologic Care") OR TX ( aged or elderly or senior or older people or geriatric or gerontologic or older adults) )

**Limiters** - Publication Year: prior to 20231231

**Expanders** - Apply equivalent subjects

**Narrow by Language:** - English

**Search modes** - Boolean/Phrase

**Scopus**

(TITLE-ABS-KEY (depression OR depressive disorder OR depressive symptoms OR major depressive disorder ) AND TITLE-ABS-KEY (integrative reminiscence OR integrative reminiscence therapy OR life review OR life story ) AND TITLE-ABS-KEY ( aged OR elderly OR senior OR older people OR geriatric OR gerontologic OR older adults OR gerontologic )
